# Use of Convalescent Plasma Therapy with Best Available Treatment (BAT) among Hospitalized COVID-19 Patients: A Multi-Center Study

**DOI:** 10.1101/2022.02.23.22271424

**Authors:** Flordeluna Mesina, Jomell Julian, Jesus Relos, Rosalio Torres, Maureen Via M. Comia, June Marie P. Ongkingco, Jimmy R. Lafavilla

## Abstract

The COVID-19 disease caused by SARS-CoV2 virus has gripped the whole world with overwhelming strain in our health system. Currently, there are no standard guidelines in its treatment but the possible benefits of convalescent plasma in limiting complications and severity of the COVID-19 disease have emerged.

**OBJECTIVE:** This study aims to determine the effectiveness and safety of using convalescent plasma in improving the clinical course of hospitalized patients diagnosed with COVID-19 disease admitted at University of Santo Tomas and Makati Medical Center.

**METHODS:** This study is a quasi-experimental (prospective analytical), and multi-center study involving 65 patients diagnosed with COVID-19 Disease who received convalescent plasma, with 65 patients who only received best available treatment serving as age-gender-matched control.

**RESULTS:** Median age of the population who received convalescent plasma was 60 years old, mostly male (68%), and manifested severe pneumonia (47%). There was noted statistically signifcant decrease between the pre-and post-treatment values of hemoglobin (p=0.04) and LDH (p=0.086). There was also statistically significant increase in platelet counts (p=0.01). WBC and PaO2 increased while ferritin and PFR decreased after convalescent plasma transfusion, however, these were not statistically significant. Length of stay and clinical outcome of those who received convalescent plasma were then compared to age-gender matched controls who only received best available treatment. There was noted statistically significant difference between length of stay (p=0.00) among those who received convalescent plasma as compared to those who did not. This was seen across severe and critically ill COVID-19 patients. There was also more mortality seen in the best available treatment alone group, but this was non-significant.

**CONCLUSIONS:** Convalescent plasma use showed no significant impact in the recovery rate and outcome of patients who received it as compared to those who did not, however, its use was proven to be safe among all patients regardless of the level of severity and clinical profile.

## INTRODUCTION

The Novel Coronavirus (COVID-19) has gripped our country with overwhelming strain in our health system. Currently, there are no standard guidelines in its treatment but the possible benefits of convalescent plasma in limiting complications and treating COVID-19 disease is being looked upon. Plasma is the liquid portion of blood containing proteins like albumin, coagulation cascade factors, complement, and a variety of antibodies or immunoglobulins and enzymes.^1^ The primary function of all immunoglobulins is the recognition and binding of specific antigenic determinants, whether soluble (including toxins), particulate, or cellular, such as pathogens. The consequences of immunoglobulin binding depend on the nature of the antigen, and at its simplest, may be the physical prevention of antigen penetration through the epithelium.^2^ This is the neutralizing effect of immunoglobulins. Secondary effects of immunoglobulin are complement activation, opsonization and antibody dependent cell mediated toxicity by different cells.

Passive antibody therapy through the use of convalescent plasma involves the administration of antibodies to a given agent to a susceptible individual for the purpose of preventing or treating an infectious disease due to that agent. In SARS-COV-2, the anticipated mechanism of action by which passive antibody therapy would act is by viral neutralization. However, it should be taken with caution since it can also trigger secondary effects mentioned above, that is important in treating patients already in the hyperinflammation state. Convalescent Plasma (CP) was used in 2013 African Ebola epidemic. A small non-randomized study in Sierra Leone revealed significant increase in survival for those treated with convalescent whole blood relative to those who used standard of treatment.^3^ Although data and study population were small, there were several studies that have documented shorter hospital stay and lower mortality in patients treated with CP than those not treated with convalescent plasma.^4-6^

The use of convalescent plasma is still an experimental therapy and is not included in the standard supportive treatment for COVID-19 disease. It is our aim to provide guidelines on the rationale and compassionate use of convalescent plasma looking into its efficacy with utmost regulation to ensure safety of both the donor and recipient.

## CONFLICT OF INTEREST

All investigators declare no conflict of interest.

## STUDY OBJECTIVES

### Primary objective

To determine the effectiveness and safety of using convalescent plasma in improving clinical course of hospitalized patients diagnosed with COVID-19. Specific objectives:

1. To describe the effect of convalescent plasma on the following clinical and laboratory parameters before and after treatment:
  a. Mean values of complete blood count (CBC), lactate dehydrogenase (LDH), serum ferritin, D-dimer, and arterial blood gas (ABG);
2. To determine the effect of convalescent plasma on the following clinical and laboratory parameters compared to historical controls that received Best Available Treatment (BAT) alone (age, gender, severity-matched controls from the same institution):
  a. Length of hospital stay
  b. Outcome;
3. To describe events of mortality in the study groups;
4. To determine the adverse effect of convalescent plasma infusion among recipients.

## METHODOLOGY

This is a quasi-experimetal (prospective analytical), multi-center study, which started its process with active recrutiment of possible donors from recovered COVID-19 patients from each institution. These possible donors were screened, and those who were selected were collected with plasma apheresis. The study population of this study included 18 years of age or older patients, hospitalised and diagnosed with COVID-19, as confirmed by positive RT-PCR for SARS-COV-2 through nasopharyngeal or oropharyngeal swab (NPS/OPS). The patients were administered with 250 to 500 ml of plasma over 1-2 hours, and were monitored for any transfusion reaction. Assessment pre-administration of the plasma was done, with descriptive data gathered, as well as, laboratory parameters such as complete blood count, serum ferritin, lactate dehydrogenase, D-dimer, and arterial blood gas. Assessment and laboratory tests were repeated 24 hours, 3 or 5 days post-transfusion of the plasma to document response.

These data of the study population who received CPT were matched based on age, gender and severity of COVID-19 pneumonia to patients who were admitted on the same time period but did not receive CPT. Reasons for non-administration of CPT include unavailability of plasma, non-consent to the procedure, and attending physician’s discretion.

## DATA ANALYSIS

Variables were expressed as n (%), and descriptive analysis of mean, median and interquartile range were used for these data. The pre-treatment and post-treatment laboratory values were compared using signed rank test. Mann-Whitney U test meanwhile was used to compare parameters from the BAT versus CPT group.

## ETHICAL CONSIDERATIONS

The investigators performed the study in accordance to the ethical principles laid down in the Declaration of Helsinki. UST Hospital Research Ethics Committee reviewed the study and granted its authority, with deviation to protocol properly documented and revised.

## RESULTS

Study was done in Hospital 1 from April 2020 to October 2020, and in Hospital 2 from October 2020 to March 2021, with total subjects of 65 patients given Convalescent Plasma Therapy (CPT).

**Table 1.** General Characteristics of the Study Population

| CHARACTERISTIC | N=65 (%) |
| --- | --- |
| AGE (in years) |  |
| 18 – 50 | 18 (28%) |
| 51 – 70 | 31 (48%) |
| 71 – 100 | 16 (24%) |
| All (median, range) | 60 years old (21 – 90) |
| SEX |  |
| Female | 21 (32%) |
| Male | 44 (68%) |
| COVID-19 PNEUMONIA SEVERITY CLASSIFICATION |  |
| Moderate | 14 (21%) |
| Severe | 31 (47%) |
| Critical | 20 (32%) |
| BASELINE (PRE-CPT) LABORATORIES |  |
| COMPLETE BLOOD COUNT |  |
| Hemoglobin (g/dl) |  |
| Hb < 12.0 | 13 (20%) |
| Hb 12.0 – 18.0 | 52 (80%) |
| Hb > 18.0 | 0 |
| Median, IR | 138 g/dl, 122 – 148.5 g/dl |
| White Blood Cell count (x10<sup>9</sup>/L) |  |
| WBC <5.0 | 12 (18%) |
| Normal 5 – 10 | 33 (50%) |
| WBC >10.0 | 20 (32%) |
| Median, IR | 7.54 x 10<sup>9</sup>/L, 5.21 – 10.3 x 10<sup>9</sup>/L |
| Platelet count (x10<sup>9</sup>/L) |  |
| <150,000 | 1 (2%) |
| 150,000-450,000 | 60 (92%) |
| >450,000 | 4 (6%) |
| Median, IR | 239,500 x 10<sup>9</sup>/L, 210.25 – 280 x 10<sup>9</sup>/L |
| INFLAMMATORY MARKERS (mean, range, SD) |  |
| LDH | 517.46 (190-3657) SD 441.48 |
| D-dimer | 4.15 (0.04-80.0) SD 12.29 |
| Ferritin | 3358.09 (289.4-40000) SD 5766.15 |
| PaO<sub>2</sub> | 125.61 (26-599.3) SD 118.96 |
| PFR | 219.01 (50-709) SD 130.98 |

IR=Interquartile range; LDH=Lactate Dehydrogenase; PaO<sub>2</sub>=Partial Pressure of Oxygen; PFR=Pao<sub>2</sub>/Fio<sub>2</sub> ratio

Median age of population is 60 years old, male predominant (68%), and manifested severe pneumonia (47%). Baseline complete blood count showed median hemoglobin of 138 g/dl, median WBC of 7.54 × 10^9^/L and median platelet count of 239,500 × 10^9^/L. Mean values of inflammatory markers are as follows: LDH 517.46 U/L (normal value: 140 – 280 U/L), D-dimer 4.15 mcg/mL (normal value: <0.4 mcg/mL), Ferritin 3358.09 mcg/L (normal value: men 24 – 336 mcg/L, women 11 – 307 mcg/L), Partial pressure of oxygen (PaO2) 125.61 mmHg (normal value: 75 – 100 mmHg), and PaO2/FiO2 ratio (PFR) 219.01 (normal value: >300 mmHg).

**Table 2.** Pre- and Post-treatment values of Set Laboratory Parameters

| PARAMETER | PRE-TREATMENT<br>VALUES<br>(mean, range, SD) | POST-TREATMENT<br>VALUES<br>(mean, range, SD) | p-value |
| --- | --- | --- | --- |
| Hemoglobin (g/dl) | 133.73 (90-171) SD 20.02 | 127.90 (59-166) SD 24.18 | 0.04 |
| WBC count | 9.73 (1.05-58.7) SD 8.08 | 10.97 (3.02-39.4) SD 6.94 | 0.16 |
| Platelet count | 263.36 (126-606) SD 11.24 | 282.77 (121-523) SD 88.31 | 0.01 |
| LDH | 517.46 (190-3657) SD 441.48 | 458.95 (198-1402) SD 29.82 | 0.086 |
| Ferritin | 3358.09 (289.4-40,000) SD 5766 | 3287.95 (506-40,000) SD 5418.67 | 0.60 |
| PaO<sub>2</sub> | 125.61 (26-599.3) SD 118.96 | 127.85 (18-618.6) SD 116.73 | 0.38 |
| PFR | 219.01 (50-709) SD 129.28 | 187.28 (23-439) SD 102.04 | 0.16 |

LDH=Lactate Dehydrogenase; PaO<sub>2</sub>=Partial Pressure of Oxygen; PFR=Pao<sub>2</sub>/Fio<sub>2</sub> ratio

There was noted statistically significant decrease between the pre- and post-treatment values of hemoglobin and LDH, and a statistically significant increase in platelet counts. WBC and PaO2 increased while ferritin and PFR decreased after treatment. However, all were not statistically significant.

Lastly, the effect of CPT to clinical parameters of length of stay and outcome was compared to historical (age-gender-matched from the same institution) controls that received best-available treatment (BAT) alone.

**Table 3.**
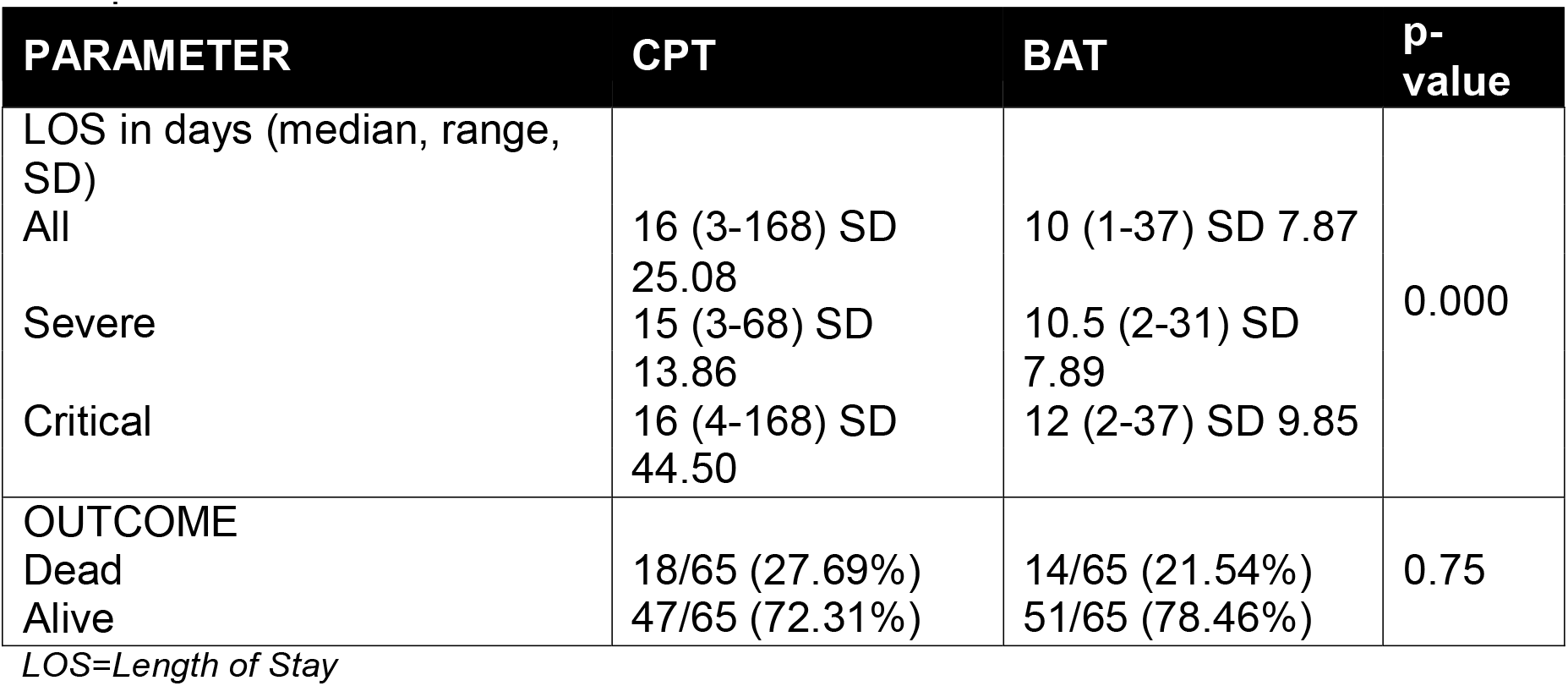
Effect of Convalescent Plasma Therapy on Set Clinical Parameters as Compared to Historical Controls Who Received BAT Alone

| PARAMETER | CPT | BAT | p-value |
| --- | --- | --- | --- |
| LOS in days (median, range, SD) |  |  |  |
| All | 16 (3-168) SD 25.08 | 10 (1-37) SD 7.87 | 0.000 |
| Severe | 15 (3-68) SD 13.86 | 10.5 (2-31) SD 7.89 |  |
| Critical | 16 (4-168) SD 44.50 | 12 (2-37) SD 9.85 |  |
| OUTCOME |  |  |  |
| Dead | 18/65 (27.69%) | 14/65 (21.54%) | 0.75 |
| Alive | 47/65 (72.31%) | 51/65 (78.46%) |  |

LOS=Length of Stay

There was noted statistically significant difference between the length of stay between those study subjects who received CPT and those who received BAT alone. This significant difference was also seen across severely- and critically-ill patients. Moreover, there were more alive than dead for those who received CPT than BAT alone, however it was not statistically significant.

## DISCUSSION

As of this writing the world has about 170,426,245 million confirmed cases, with 3,548,268 million deaths, of COVID-19 infection since it was first reported last December 2019 in Wuhan, Hubei Province, China.^8^ It became one of the worst pandemic in the history of the world, together with HIV/AIDS, and it is still causing physical, emotional and economical toll worldwide.

No single best treatment for COVID-19 infection has been identified yet. Some of the available treatments being used are Dexamethasone shown to have mortality benefits for patients in respiratory support, Remdesivir that was approved by US FDA last October 2020 and was shown to modestly speed up recovery time, and anticoagulants such as enoxaparin as thromboprophylaxis. In August 2020, US FDA issued an emergency use authorization (EUA) for Convalescent Plasma Therapy (CPT) for patients hospitalized with COVID-19 infection. Earlier studies showed promising results of mortality benefits, however no control group was utilized.^9^ Nonetheless, a randomized double-blind and placebo-controlled trial published in The New England Journal of Medicine last November 2020 showed that patients who received CPT within three days were 48% less likely to develop severe COVID-19 infection compared to those who received placebo, however this finding was not statistically significant.^9,10^

Locally, the Philippines has a total of 1,230,301 confirmed COVID-19 cases since its first confirmed case last January 2020.^11^ Local hospitals, especially major tertiary ones in Metro Manila were able to use CPT adjunctively. UP-PGH is one of three major government institutions assigned to accommodate COVID-19 patients with a total of 4,540 cases since February 2020, and has an undergoing CPT study.^12^ This multi-center study was not far behind in taking off, with an initial number of 523 admitted cases when the use of CPT started last April 2020.

Only sixty-five patients who received CPT in this multi-center study were eligible after narrowing down with the inclusion/exclusion criteria. Most were of 51 – 70 years of age (48%), with median age of 60, and predominantly males (68%). These findings are in line with those from CPT studies done in Argentina (median age of 62 years, 67.6% men),^10^ the Netherlands (median age of 63 years),^13^ the PLACID trial of India (75.3% males),^14^ China (median age of 61.5 years, 83.3% males^15^; 58.3% males^16^) and the US (64% males).^16^ One study in China^4^ found that most of elderly patients (>60 years of age) presented with severe and critical types of COVID-19 infection, and identified age and gender with morbidity as risk factors. They also noted that the elderly have worse survival outcome compared to younger patients, with poor nutritional status and immune functions as probable reasons for this. In this multicenter study, most of those who belong to the elderly age presented with severe to critical types of COVID-19 infection, thus an indication to use CPT. The Center for Disease Control and Prevention (CDC) also identified that the overall COVID-19 case-fatality ratio is higher among men than women, both in China and the US, with biological, psychosocial and behavioral factors might be affecting these findings.^18^ However, WHO recognized the disparity of available sex- and age-disaggregated data hampering the analysis of its association to COVID-19 infection and possible implications.

Most of the patients who received CPT manifested severe COVID pneumonia (47%), which clinically presents with fever, tachypnea of >30 breaths per minute or spO2 of <93% at room air, and sepsis or septic shock. As mentioned above, more severe type of COVID-19 infection would indicate use of CPT. However, previous studies showed no clinical significance in outcomes of patients with moderate or even severe or life-threatening disease who used CPT.^11^ The median day from admission to convalescent plasma therapy for this multi-center study was 3 days, and earlier studies showed effectiveness of CPT if given within three days,^10^ however, some outcomes were not also statistically significant for this multicenter study, as will be discussed further later. The improvement shown in some of the outcomes after CPT in this multi-center study was probably compounded by the use of other treatment modalities such as proning-positioning, respiratory support and hemoperfusion, and medications such as antibiotics like Meropenem (43%), Azithromycin (39%), Levofloxacin and Piperacillin and Tazobactam (17%), Cefepime and Tigecycline (13%), and Ceftriaxone, Doxycycline, Metronidazole and Moxifloxacin (8%), anti-viral Remdesivir (73%), anti-fungal Fluconazole (13%), MAB Tocilizumab (21%), thromboprophylaxis Enoxaparin (65%), and Dexamethasone (78%), and cannot be solely be attributed to CPT alone. However in the study by Simonovich et al., CPT in addition to standard treatment with severe pneumonia still did not reduce mortality or improved clinical outcomes at day 30 compared to placebo.^10^

Infection such as with COVID-19, are long been known to affect not only clinical parameters of the patient but laboratory parameters as well. More commonly, acute infection would often lead to anemia, mainly due to hepcidin-mediated iron-restricted erythropoiesis and erythrocyte production suppression by cytokines thru its act on erythroid progenitors;^19^ leukocytosis which is a more common response of a normal bone marrow to inflammation or infection,^20^ as infection or inflammation is a stimuli that would lead to increased production of granulocytic marrow and release of fully matured neutrophils stored in the bone marrow or marginated along the endothelium of the vasculature;^21^ and thrombocytosis, as platelets and its activation can regulate innate and adaptive immune responses, with host inflammatory responses resulting to release of platelet activating mediators, and sometimes thrombocytopenia, thru consumption.^22^ In a study by Terpos et al., they found out that during incubation period and early phase of COVID-19 infection, peripheral leukocyte and lymphocyte counts are slightly reduced, with pronounced lymphopenia only evident after 7 to 14 from onset of symptoms.^23^ This is compounded with increase of inflammatory mediators and cytokines, and can even be characterized as cytokine storm. Aside from lymphopenia, there was also noted thrombocytopenia in COVID-19 patients, and this is true for other studies from Guan et al.^24^ and Fan et al.^25^ Hemoglobin and red blood cell meanwhile were significantly lower in severe or critically ill COVID-19 patients as compared to regular ones.^26^ In this multi-center study, median values and interquartile ranges were all within normal range [Hemoglobin: 138 g/dl, 122 – 148.5 g/dl; WBC: 7.54 × 10^9^/L, 5.21 – 10.3 × 10^9^/L; Platelet count: 239,500 ×10^9^/L, 210.25 – 280 × 10^9^/L]. This might be explained by the timing of admission and initial evaluations of these patients as mostly were at the beginning of their illness. Upon intervention of CPT, although there was noted significant decrease in the mean hemoglobin value, it is still within the normal. Leukopenia and thrombocytopenia were also not observed after CPT intervention, with significant increase in platelets noted after therapy, and values still within the normal range. On previous CPT studies, only improvement with lymphocyte counts upon CPT was frequently reported.^27,28^ Thus, the effect on hematological indices such hemoglobin and platelet counts as reported in this multi-center study could be further investigated on future CPT studies.

Inflammatory markers have also been associated with severity of COVID-19 infection, however findings remain inconclusive.^29^ These inflammatory markers, or acute phase reactants, are produced in the liver during inflammatory states, and can be classified as positive acute phase reactants (i.e. procalcitonin, CRP, ferritin, fibrinogen, hepcidin, and serum amyloid), which are upregulated during inflammation, or negative acute phase reactants (i.e. albumin, prealbumin, transferrin, retinol-binding protein, and anti-thrombin), which are downregulated during inflammation.^30^ Lactate dehydrogenase is also considered as an inflammatory marker, and is a general indicator of tissue damage, especially in the lungs.^31^ In this multi-center study, inflammatory markers ferritin and LDH were tested, and both mean values are found to be elevated [LDH: 517.46 (190-3657) SD 441.48, Ferritin: 3358.09 (289.4-40,000) SD 5766], and these findings are consistent with other studies.^29,32^ After intervention with CPT, LDH and ferritin were both found to be decreased, but only LDH reduction was noted to be significant. In small studies involving 4 to 8 patients who presented from mild to critical COVID-19 infection, there was noted decrease in LDH levels after CPT.^33^ But a much bigger study involving 233 patients who have either moderate or severe COVID-19 infection, found that there was no significant improvement of LDH levels after CPT.^34^ Ferritin, meanwhile also has conflicting findings. Simonovich et al. (2021)^10^ found no difference on ferritin levels between patient groups at day 14, while the study from the US by Salazar et al., found decreasing trends of ferritin levels by day 3.^35^

COVID-19 is a multi-systemic infection that mainly involves the lungs, and approximately 14% of patients require hospitalization and oxygen support, and 5% require ICU admission. Severe cases are usually complicated with Acute Respiratory Distress Syndrome (ARDS), which causes oxygenation impairment [i.e. 200 mmHg < PaO2/FiO2 ≤ 300 mmHg with PEEP or CPAP ≥5 cmH2O or non-ventilated for Mild; 100 mmHg < PaO2/FiO2 ≤ 200 mmHg with PEEP ≥5 cmH2O or non-ventilated for Moderate; and PaO2/FiO2 <100 mmHg with PEEP >5 cmH2O or nonventilated for Severe ARDS; and if PaO2 is not available, SpO2/FiO2 ≤315 suggests ARDS, including those who are non-ventilated]. This is due to the nature of COVID-19 infection wherein there would be elicited surfactant changes that causes its downregulation, changes in cell-matrix adhesion which could lead to epithelial cell dysfunction and changes in core ECM which leads to lung tissue damage.^36^ In this multi-center study, mean value of PaO2 was not found to be decreased, however PFR was noted to be impaired. On CPT intervention, both were found to have not improved significantly. However in previous CPT studies, they reported significant improvement on both PaO2 and PFR after CPT intervention.^5,37-39^

In the study of Simonovich et al, they compared the clinical status of COVID-19 patients who received CPT from those who received only placebo, and no significant difference was noted on the clinical outcomes of the patients after day 30.^10^ Mortality at day 30 was noted to be higher on placebo group with - 0.46 risk difference. In this multi-center study, length of stay was significantly longer on those who received CPT than those who only received BAT, even in those who were severely or critically ill. However, the BAT group has less number of mortality as compared to the CPT group, although this comparison was non-significant. Both of these findings may be explained by several observations:

1. Length of stay was longer for the CPT group as the intervention may had prevented early mortality; but the worse presentation of the illness for the CPT group causing longer hospital days and mortality later on, can also be a reason; or
2. The better presentation of the illness for the BAT group can explain the shorter length of stay, and less number of mortality for the BAT group.

These conflicting findings are in line with several CPT studies that found hospital stay and mortality benefits with CPT ^35,40-42^ and those that found no significant difference.^16^ In a local study done in Cardinal Santos Medical Center,^46^ they also found no difference in mortality and length of hospital stay in patients given with CPT as compared to controls, however they noted improvement in inflammatory markers and pulmonary status in CPT recipients.

In a retrospective analysis done among 77 COVID-19 deaths by Wang et al. (2020), 72.7% of the study population had co-morbidities, with male predominance, mean age of 71±13 and mean survival time of 17.4±8.4 days.

There was noted lymphopenia and increase in inflammatory markers among these patients. The most common causes of death were acute respiratory failure (ARF) and sepsis. However, the proportion of younger deaths was higher in males, and MI mainly caused elderly deaths.

More than twenty-seven percent among the study participants in this multi-center study died during their hospital stay. Mostly presented with severe (44%) and critical (51%) infection, however, age among these patients varied from 39 to 81 years of age, with predominance of the male population (55%). Length of stay also varied from as short as 3 days to as long as 168 days. Duration from admission to CPT also differed from day 0 to day 12. Mostly died primarily due to ARF or MI. However, other co-factors that might lead to demise were not investigated, and clinical and laboratory parameters were not compared between those who died and those who were alive, before and after CPT. This could be a good recommendation for next CPT studies.

Like any other blood transfusion, the most common adverse reaction observed in CPT is transfusion-related events such as chills, fever, anaphylactic reactions, TRALI, circulatory overload and hemolysis. Generally, no serious adverse events during CPT were observed in this multi-center study. Only one patient experienced mild hypersensitivity reaction, and was immediately managed with conventional anti-pyretic and anti-histamine.

## CONCLUSION

In this study, convalescent plasma may have shown no significant impact in the recovery rate and outcome compared to patients who have not received convalescent plasma therapy, but its administration was proven to be safe among all patients regardless of the level of severity and clinical profile.

## STUDY LIMITATIONS

The study did not look into the level of viral neutralization of immunoglobulins from the convalescent plasma. Ideally, quantitative antibody titer of donors should be done prior to collecting plasma but this test was not locally available at the time of the study. Instead, the study used the qualitative method of antibody identification or a surrogate marker was used that may approximate antibody titer. Determination of the effect of CPT to mean ROX index, and imaging were also not pursued due to inconsistent availability of these values/tests across the sample population.

## RECOMMENDATIONS

A sub-analysis may benefit this multicenter study for further clinical significance. Evaluation for day of illness to the day of convalescent plasma administration and antibody titer measurements may be further investigated. Factors for the length of stay and outcome such as hospital-acquired infection and comorbidities may also be explored.

## Data Availability

All data produced in the present study are available upon reasonable request to the authors.

## Notes

### Competing Interest Statement

The authors have declared no competing interest.

### Funding Statement

This study received a partial assistance from the Philippine College of Hematology and Transfusion Medicine(PCHTM).

### Author Declarations

Research ethics committee/IRB of University of Santo Tomas Hospital and Makati Medical Center gave ethical approval for this work.

